# Neuroimaging Correlates of Compulsive Sexual Behavior and Problematic Pornography Use: A Systematic Review and Coordinate-Based Meta-Analysis Protocol

**DOI:** 10.1101/2025.06.12.25329499

**Authors:** Yuan Feng, Zhonghua Wei

## Abstract

Compulsive sexual behavior disorder (CSBD) and problematic pornography use (PPU) have garnered increasing attention as clinically significant conditions, potentially falling within the spectrum of behavioral addictions or impulse control disorders. However, neuroimaging findings to date have been inconsistent and scattered across studies, limiting our understanding of the neural mechanisms underlying these conditions. This protocol outlines a systematic review and coordinate-based meta-analysis (CBMA) designed to synthesize structural and functional neuroimaging results related to CSBD and PPU. By aggregating stereotactic coordinates from eligible studies, we aim to identify convergent patterns of brain alterations, particularly in regions implicated in reward processing, executive control, and salience attribution. The inclusion of both resting-state and task-based imaging data is expected to improve statistical power and facilitate a more integrative characterization of the neural correlates of CSBD and PPU. Findings from this review may help clarify the extent to which these conditions share neurobiological features with other addictive behaviors. This will advance theoretical models of dysregulated sexual behavior and inform future diagnostic and therapeutic strategies.

## 1. Introduction

### 1.1 Background

Compulsive sexual behavior disorder (CSBD) and problematic pornography use (PPU) have attracted growing attention in recent years and are increasingly regarded as potential forms of behavioral addiction or impulse control disorders. CSBD is defined as a persistent failure to control intense, repetitive sexual impulses or urges, leading to repetitive sexual behavior that causes significant distress or impairment[1]. PPU is often considered a specific manifestation of CSBD with a predominant focus on online sexual content[2]. Estimated prevalence of high-risk CSBD ranges from 0–5.5% in women and 4.2–7% in men[3]. Although CSBD has been increasingly studied and formally recognized in diagnostic systems such as the ICD-11, substantial controversy remains regarding its conceptualization and clinical management[4]. As a psychiatric condition rooted in fundamental human drives, CSBD remains comparatively under-investigated relative to other mental disorders.

In parallel with its growing clinical relevance, an increasing number of neuroimaging studies have sought to elucidate the neural underpinnings of CSBD and PPU. Previous neuroimaging studies have reported increased activation in the ventral striatum and orbitofrontal cortex, along with enhanced functional connectivity between the inferior frontal gyrus and widespread cortical regions[5,6]. Structurally, individuals with CSBD have shown reduced cortical surface area in the right posterior cingulate cortex, negatively associated with symptom severity[7]. Additionally, decreased fractional anisotropy has been observed in the superior corona radiata, internal capsule, cerebellar pathways, and occipital white matter[8]. These investigations have reported alterations in brain regions associated with reward processing, cognitive control, and salience detection, such as the ventral striatum, prefrontal cortex, and cingulate cortex. However, the findings remain inconsistent across studies, partly due to variations in diagnostic criteria, sample characteristics, and imaging paradigms. Although diagnostic heterogeneity remains a challenge, coordinate-based meta-analyses can help identify spatially consistent neural correlates despite such variability. While some narrative reviews have summarized existing neuroimaging studies on CSBD and PPU, no coordinate-based meta-analytic efforts have been undertaken to identify convergent neural signatures.

Despite the growing number of individual neuroimaging studies, no prior review has specifically focused on summarizing brain imaging findings in CSBD or PPU, and coordinate-based meta-analytic synthesis is entirely lacking in this field. This represents a critical gap in the literature, given the clinical and theoretical relevance of identifying reliable neural correlates. Without such a synthesis, it remains difficult to determine whether these behaviors share neurobiological features with substance use or other behavioral addictions.

This review aims to fill this gap by aggregating stereotactic coordinate data from multiple neuroimaging modalities—including structural and functional MRI—to identify spatially convergent alterations across studies. By doing so, we aim not only to address between-study heterogeneity but also to highlight the brain regions most consistently implicated across modalities and paradigms. These findings may inform diagnostic classification frameworks and support the development of targeted, neurobiologically informed interventions. To address these gaps, we formulated the following review objectives and questions.

### 1.2 Objectives and Review Questions

#### Objective

This review aims to systematically identify and synthesize neuroimaging findings related to CSBD and PPU. Specifically, we aim to determine whether consistent patterns of brain structural or functional alterations exist across studies, and whether these patterns overlap with those implicated in other addictive or impulse control disorders.

By characterizing the neural correlates of CSBD and PPU, this review seeks to advance our understanding of their underlying neurobiological mechanisms. Such insights may facilitate differentiation from other behavioral addictions, inform ongoing debates surrounding their nosological classification, and provide a theoretical foundation for the development of targeted interventions aimed at modulating specific brain circuits.

#### Review Questions

1. What are the consistent neural correlates of compulsive sexual behavior and problematic pornography use based on structural and functional neuroimaging findings?
2. Do individuals with CSBD or PPU show altered activity or structure in brain regions involved in reward processing, cognitive control, or emotional regulation?
3. Are the identified neural patterns similar to those observed in other behavioral or substance addictions?
4. How do participant characteristics (e.g., age, gender) or study parameters (e.g., imaging modality, task type) moderate the observed neural findings?

### 1.3 Outcomes

The primary outcome of this review is the identification of brain regions that show consistent structural or functional differences between individuals with CSBD or PPU and healthy controls. Specifically, we will extract peak stereotactic coordinates (in MNI or Talairach space) of altered brain activity or structure as reported in eligible neuroimaging studies. These coordinates will be used in a coordinate-based meta-analysis to determine convergent neural correlates of CSBD and PPU.

Secondary outcomes will include alterations in functional connectivity patterns, the involvement of specific brain networks (e.g., the reward system, executive control network), and the role of task paradigms (e.g., cue reactivity, inhibitory control) as potential moderators of neural activity. Where available, we will also examine the influence of participant characteristics (e.g., age, gender) and imaging modality (e.g., functional vs. structural MRI) on the observed neural findings.

## 2. Methods

We will use Neurosynth Compose (https://neurosynth.org/compose/) as the primary data source to identify relevant neuroimaging studies. Neurosynth Compose is an advanced platform that integrates a large corpus of brain imaging literature with automated tools for coordinate-based meta-analysis. To ensure comprehensive and precise retrieval, we will combine automated keyword-based searches with manual screening and verification of eligible studies.

### 2.1 Information sources and search strategy

We will input a comprehensive set of search terms related to compulsive sexual behavior and pornography use into the Neurosynth Compose platform, including: “Compulsive Sexual Behavior” OR “Compulsive Sexual Behavior Disorder” OR “CSBD” OR “Sex Addiction” OR “Pornography Addiction” OR “Problematic Pornography Use” OR “Porn Use Disorder” OR “Cybersex Addiction” OR “Hypersexuality” OR “Sexual Compulsivity” OR “Sexual Impulsivity” OR “Sexual Craving” OR “Sexual Cue Reactivity” OR “Excessive Sexual Behavior” OR “Problematic Online Sexual Behavior” OR “Sexual Dysregulation” OR “Sexual Obsession” OR “Online Sex Addiction” OR “Erotic Stimuli” OR “Sexual Stimuli” OR “Sexual Arousal”.

Neurosynth Compose will be used to identify relevant studies indexed in the Neurosynth database that report stereotactic coordinates (in MNI or Talairach space) of brain activations or structural alterations associated with CSBD or PPU. All identified records will be manually screened to ensure they meet the eligibility criteria (see Section 2.2), and duplicate entries will be removed. Only peer-reviewed articles published in English will be considered. In addition, we will manually search reference lists of included studies and relevant reviews to identify any additional eligible publications.

### 2.2 Study selection

All search results retrieved from the Neurosynth Compose platform will be processed according to its built-in PRISMA-based workflow. The platform provides structured steps for de-duplication, initial title and abstract screening, and full-text selection, aligned with the PRISMA 2020 guidelines. Two reviewers will independently screen the titles and abstracts of all retrieved studies. Full texts of potentially relevant articles will then be examined to determine eligibility based on predefined inclusion and exclusion criteria (see Sections 2.2.1 and 2.2.2). Discrepancies between reviewers will be resolved through discussion, and a third reviewer will be involved if necessary.

The selection process will be documented and visualized using a PRISMA 2020 flow diagram generated through Neurosynth Compose. Additional records identified through manual search of reference lists or external databases (if any) will be incorporated and documented in the PRISMA diagram.

#### 2.2.1 Inclusion criteria

(1) Participants are adults (aged 18 years or older) diagnosed with CSBD, PPU, hypersexuality, or self-reported sexual behavior addiction. (2) Alternatively, studies include individuals who score above validated thresholds on standardized measures of compulsive sexual behavior or pornography use (e.g., SAST-R, CSBD-19, Brief Pornography Screen). (3) The study includes a healthy control group or a between-group comparison relevant to CSBD or PPU.(4) The study employs neuroimaging techniques, including functional MRI, structural MRI, diffusion tensor imaging (DTI), or positron emission tomography (PET), to examine brain structure or function in relation to CSBD or PPU. (5) The study reports stereotactic coordinates (in MNI or Talairach space) from between-group comparisons that can be used for coordinate-based meta-analysis. (6) The full text is available in English and published in a peer-reviewed journal.

#### 2.2.2 Exclusion criteria

(1) The study includes only healthy participants or focuses solely on experimental models without a comparison to an at-risk or affected group. (2) The study involves individuals with sexual dysfunction due to unrelated medical, neurological, or hormonal conditions (e.g., stroke, Parkinson’s disease, menopause). (3) The disorder of interest (CSBD or PPU) is not the primary focus of the study (e.g., it is treated only as a comorbid condition or secondary outcome). (4) The study does not report stereotactic coordinates (in standard space), or such data are unavailable for analysis. (5) The study involves non-human subjects. (6) Studies that solely focus on machine learning-based classification or prediction, without reporting interpretable group-level neuroimaging differences relevant to CSBD or PPU. (7) The publication is a review, meta-analysis, single case report, conference abstract, or other non-primary research format.

### 2.3 Data extraction

Key study-level data will be extracted using a standardized data extraction form developed for this review. The following information will be collected from each included study: (1) bibliographic details (authors, year, journal), (2) sample characteristics (e.g., sample size, age, sex), (3) diagnostic criteria or assessment tools used (e.g., clinical diagnosis, SAST-R, CSBD-19), (4) imaging modality and acquisition parameters (e.g., fMRI, structural MRI, DTI), (5) task paradigm (e.g., cue reactivity, resting-state, inhibitory control), (6) reported group-level brain activation or structural differences, and (7) stereotactic coordinates (in MNI or Talairach space), if available.

Coordinate data will be extracted for eligible between-group comparisons and used in coordinate-based meta-analysis. All data will be extracted independently by two reviewers, with discrepancies resolved through discussion or consultation with a third reviewer if needed.

### 2.4 Risk of bias assessment

Given the methodological heterogeneity of neuroimaging studies, traditional tools for risk of bias assessment may not fully capture the quality dimensions relevant to coordinate-based analyses. Therefore, we will use a modified version of the Joanna Briggs Institute (JBI) Critical Appraisal Checklist for Analytical Cross Sectional Studies, adapted for neuroimaging research. The checklist will focus on sample characteristics, diagnostic clarity, imaging methodology, and the transparency of reported results. This evaluation will inform sensitivity analyses but will not be used to exclude studies.

### 2.5 Data synthesis

If sufficient coordinate data are available, we will conduct a coordinate-based meta-analysis (CBMA) using the MKDAChi2 algorithm implemented in Neurosynth Compose, supported by the NiMARE framework. This method aggregates stereotactic coordinates across studies to assess the spatial consistency of reported brain alterations. Statistical significance will be determined using false discovery rate (FDR) correction with a threshold of q < 0.05 applied to the uniformity test map.

Given the relatively small number of neuroimaging studies available in this emerging research domain, FDR correction is selected over family-wise error (FWE) correction to increase statistical sensitivity while still controlling for multiple comparisons. This approach aligns with the default methodology of Neurosynth Compose and is appropriate for conditions where the expected number of included studies is limited.

In the event of insufficient coordinate availability, a qualitative synthesis will be performed to summarize reported brain regions, network-level alterations, and task paradigms.

Sensitivity analyses will be conducted, where feasible, to evaluate the robustness of the meta-analytic findings. These may include leave-one-out analyses and re-analyses excluding studies assessed as high risk of bias. The feasibility of such analyses will depend on the number of eligible studies included. We will also explore whether diagnostic method (e.g., clinical diagnosis vs. self-report) or imaging modality influences the convergence of activation patterns.

### 2.6 Visualization and Reporting

All significant clusters identified through coordinate-based meta-analysis will be visualized using standard neuroimaging tools. We plan to use DPABI (v8.1) and BrainNet Viewer to map and render brain regions onto a standard MNI template. Peak MNI coordinates of each significant cluster will be extracted and labeled using the AAL3 atlas with the aid of xjView. Surface and volume visualizations may also be generated using Surf Ice or MRIcroGL, depending on visualization needs. The final visualization tools will be selected based on the clarity, compatibility, and visual quality of outputs for the identified results.

## 3. Discussion

This systematic review and coordinate-based meta-analysis will provide a comprehensive and up-to-date synthesis of neuroimaging findings related to CSBD and PPU. By systematically aggregating reported activation and structural differences across studies, we aim to identify consistent neural correlates associated with these conditions. Although many studies have suggested that the neurobiological mechanisms of CSBD and PPU are closely related to addiction and impulse control[9,10], the core neural targets identified across studies remain inconsistent and fragmented. This lack of convergence hampers both mechanistic understanding and the development of targeted neuromodulation interventions.

This study employs CBMA to identify spatially consistent neural correlates of CSBD and PPU across heterogeneous studies. By synthesizing reported stereotactic coordinates, CBMA enhances reproducibility and enables robust localization of key brain regions, despite variations in methodology or diagnostic criteria[11]. Moreover, by integrating structural and functional imaging findings—across both resting-state and task-based paradigms—this multimodal approach improves statistical power and offers a more comprehensive understanding of the neural mechanisms underlying compulsive sexual behavior. To our knowledge, this is the first coordinate-based meta-analysis to quantitatively synthesize neuroimaging findings in CSBD and PPU, offering a first quantitative synthesis to guide both research and clinical translation in this emerging field.

Previous research has proposed that brain, cognition, and behavior function as an integrated system in the regulation of compulsive behaviors[12]. Neuroimaging techniques, with their high spatial resolution, serve as powerful tools to probe the neural substrates underlying these behaviors. In recent years, the accumulation of diverse imaging modalities—including gray matter morphometry, white matter tract integrity, resting-state connectivity, and task-based activation—offers a rich dataset for exploration. Leveraging such multimodal information allows for complementary insights and may increase the likelihood of identifying reliable neural correlates across studies[13].

This study is expected to identify consistent neuroanatomical and functional alterations in CSBD and PPU, particularly in regions related to reward processing, impulse control, and salience attribution (e.g., ventral striatum, prefrontal cortex, cingulate cortex). These findings may clarify whether CSBD and PPU share neural features with other behavioral or substance addictions, contributing to ongoing nosological debates[14]. Theoretically, this work will help elucidate how dysregulated sexual behavior may stem from imbalances among brain systems involved in motivation, self-regulation, and emotion, offering a clearer neural model for future research and hypothesis generation in clinical neuroscience.

By identifying convergent neurobiological alterations in individuals with CSBD and PPU, this review may contribute to the refinement of diagnostic classification systems such as the ICD and DSM[15]. Specifically, the results may support the inclusion of neural biomarkers that distinguish CSBD and PPU from normative sexual behavior or other psychiatric conditions. From a clinical standpoint, the delineation of core brain circuits could inform the development of targeted neuromodulation strategies (e.g., TMS, tDCS) and tailored psychotherapeutic interventions[16]. Furthermore, better neurobiological understanding may aid in early identification, destigmatization, and the formulation of evidence-based public health responses to problematic sexual behaviors.

## Limitations and Future Directions

However, some limitations should be noted. First, the available literature may be sparse and heterogeneous, limiting the statistical power of the meta-analysis. Second, inclusion is restricted to studies reporting stereotactic coordinates, which may exclude relevant but non-standardized findings. Finally, although we will attempt to explore potential moderators, limited reporting across primary studies may hinder subgroup analyses. Despite these constraints, the planned review will serve as an important step toward consolidating current neuroimaging evidence and guiding future research.

Future studies should standardize diagnostic criteria, ensure gender-balanced samples, and adopt harmonized imaging paradigms. Sharing unthresholded statistical maps (e.g., via NeuroVault) will facilitate more precise image-based meta-analyses. Longitudinal and interventional research is also needed to clarify causal links between brain alterations and CSBD/PPU symptoms.

## Ethics and Dissemination

This study involves analysis of published data and does not require ethical approval. The findings will be disseminated through medRxiv preprint publication and submission to a peer-reviewed journal.

## PROSPERO Registration

CRD420251071407

## Data Availability

This study is a protocol and does not involve original data collection. Coordinate data and study-level variables will be extracted from peer-reviewed publications and supplementary materials, where available.

